# Blood-based PhenoAge may reflect disease burden rather than biological aging in cancer

**DOI:** 10.64898/2026.10.15.26365671

**Authors:** Anish K. Simhal, Jasme Lee, Kay See Tan, Patrick J. McCormick, Gregory W. Fischer, Joseph R. Scarpa, Guillermo Sapiro, Joshua S. Mincer

## Abstract

PhenoAge, a blood-based biological aging clock, is associated with cancer outcomes, a finding taken as evidence that biological aging shapes prognosis. In 3,067 patients undergoing primary tumor resection, PhenoAge acceleration rose with tumor stage. One or two of its eight blood components, differing by cancer type, carried the prognostic signal, and replacing its general-population weights with equal weights did not weaken it. In cancer, PhenoAge may reflect disease burden rather than biological aging.

## Main

Chronological age is central to cancer risk stratification. Age alone, however, cannot explain why two people of the same age differ so markedly in cancer susceptibility and prognosis. In aging research more generally, the same gap has motivated the idea of “biological age” and a proliferation of “clocks” built to measure it^1^. One such clock, Phenotypic Age (PhenoAge)^1,2^, a blood-based estimator trained to predict mortality in the U.S. National Health and Nutrition Examination Survey (NHANES), has been adopted widely in medical research^3–6^, including cancer. Higher PhenoAge and higher PhenoAge acceleration have been linked to cancer incidence and death^7,8^ and proposed as an explanation for the rise in early-onset cancers^3^.

Cancer is characterized not only by malignant growth but also by systemic inflammation, immune dysregulation, nutritional depletion and organ dysfunction^11^. These processes are reflected in routine laboratory tests, which are ordered frequently in cancer care. PhenoAge is a weighted sum of these same laboratory values, its components, with weights fitted to predict mortality, including death from cancer, in the general population. In a patient with an active tumor, a high PhenoAge could reflect either biological aging or pathophysiology associated with the malignancy and related therapies. While both interpretations may be related to poor outcomes, they have different implications and may call for different interventions. If PhenoAge measures aging in these patients, its population-derived weights should carry prognostic information, and the full composite should outperform its parts.

We retrospectively examined treatment-naive patients undergoing curative primary tumor resection at Memorial Sloan Kettering Cancer Center (MSK), drawn from four cohorts associated with previously published studies: colon adenocarcinoma (COAD, n=577)^12^, renal cell carcinoma (RCC, n=1,724)^13^, lung adenocarcinoma (LUAD, n=241)^14^ and triple-negative breast cancer (TNBC, n=525)^15,16^ (Supplementary Table S1). PhenoAge was computed from blood drawn prior to surgery. Because C-reactive protein (CRP) is rarely measured before surgery, we used an eight-component PhenoAge^15,16^, with the CRP term omitted and the weights re-estimated in NHANES III^17,18^ (Supplementary Table S2). For comparison, applying the original nine-component formula with missing CRP set to a population constant gave near-identical rankings (Spearman ρ > 0.96) and concordance indexes (C-indexes) within 0.02 on both endpoints in every cohort (Supplementary Table S2). We tested associations with overall and recurrence-specific survival, measured from surgery.

### PhenoAge acceleration predicts outcome in all four cancers

PhenoAge acceleration is the difference, in years, between a patient’s PhenoAge and chronological age. Higher acceleration was associated with shorter overall survival in every cohort, with a hazard ratio per standard deviation (SD) of 1.66 in COAD, 1.57 in RCC, 1.28 in LUAD and 1.17 in TNBC, and with shorter recurrence-specific survival (1.39, 1.39, 1.24 and 1.13). The 95% interval excluded 1 in every case except LUAD overall survival (22 deaths) and TNBC recurrence-specific survival. Kaplan– Meier analysis illustrates these results: tertiles of acceleration separated overall-survival curves in COAD, RCC and LUAD (Fig. 1). This association agrees with previous reports, which interpreted it as evidence that accelerated biological aging is associated with shorter survival in patients with cancer. However, those reports did not examine the contribution of PhenoAge’s components, as critically needed to validate if it represents aging or disease burden.

**Figure 1.**
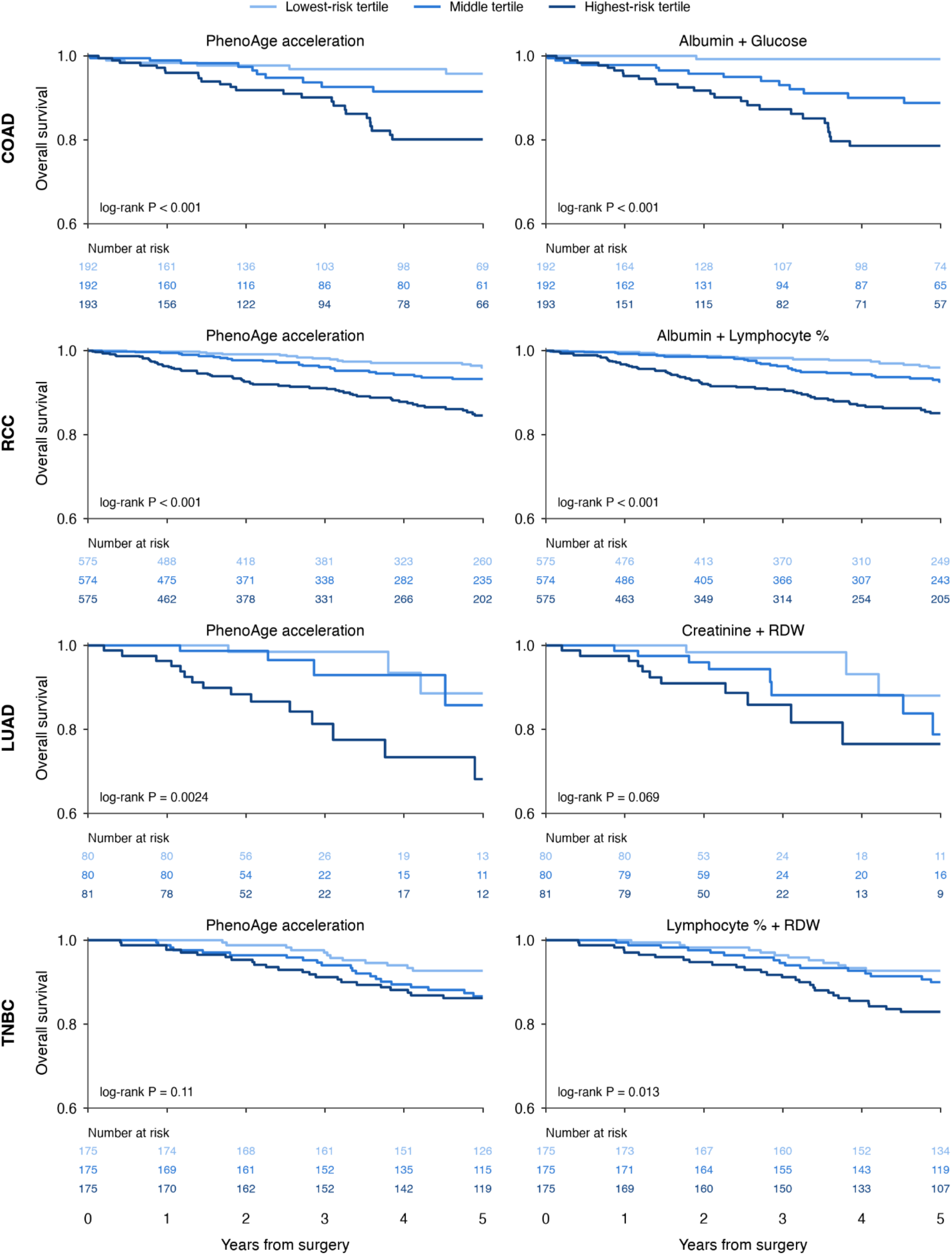
Overall survival by tertiles of PhenoAge acceleration and of a representative two-component composite. Kaplan–Meier overall survival by tertiles of PhenoAge acceleration (left column) and by tertiles of a two-component composite (right column), one row per cancer: COAD (n=577, 41 deaths within five years), RCC (n=1,724, 101), LUAD (n=241, 22), and TNBC (n=525, 56). The composite is the linear predictor of a Cox model fitted on a representative pair of components for that cancer, both standardized: albumin + glucose (COAD), albumin + lymphocyte percentage (RCC), creatinine + RDW (LUAD), lymphocyte percentage + RDW (TNBC). Tertiles are oriented so that tertile 3 is the highest-risk group. Follow-up is censored at five years so that the four cancers share one axis; patients still alive at five years remain in the analysis. P values are unstratified log-rank tests on the same five-year follow-up; numbers at risk are given beneath each panel; the survival axis is truncated at 0.6 in all panels. The curves are descriptive. Each pair is built on the single most prognostic component in that cancer (Fig. 2b); in LUAD and TNBC two or three pairs discriminate equally well, and the one shown is representative. COAD, colon adenocarcinoma; LUAD, lung adenocarcinoma; RCC, renal cell carcinoma; RDW, red cell distribution width; TNBC, triple-negative breast cancer.

### PhenoAge acceleration rises with tumor burden and adds little beyond known clinicopathologic factors

Acceleration was higher in patients whose primary tumor was more advanced at pathology (Fig. 2a): Spearman ρ with T stage was 0.20 in COAD (P < 0.001), 0.14 in RCC (P < 0.001) and 0.15 in LUAD (P = 0.023), but 0.06 in TNBC (P = 0.14), where nearly every tumor was T1 or T2 (519 of 525) and there was little stage variation to track. T stage is a coarse measure of disease burden, yet in the patients with an ordinal T stage, adjusting for it alone reduced the PhenoAge acceleration hazard ratio for overall survival from 1.67 to 1.56 in COAD, 1.57 to 1.46 in RCC and 1.28 to 1.25 in LUAD, a 10–17% reduction in the log hazard ratio; in TNBC, where the association was weakest, it was unchanged (1.17 to 1.18).

**Figure 2.**
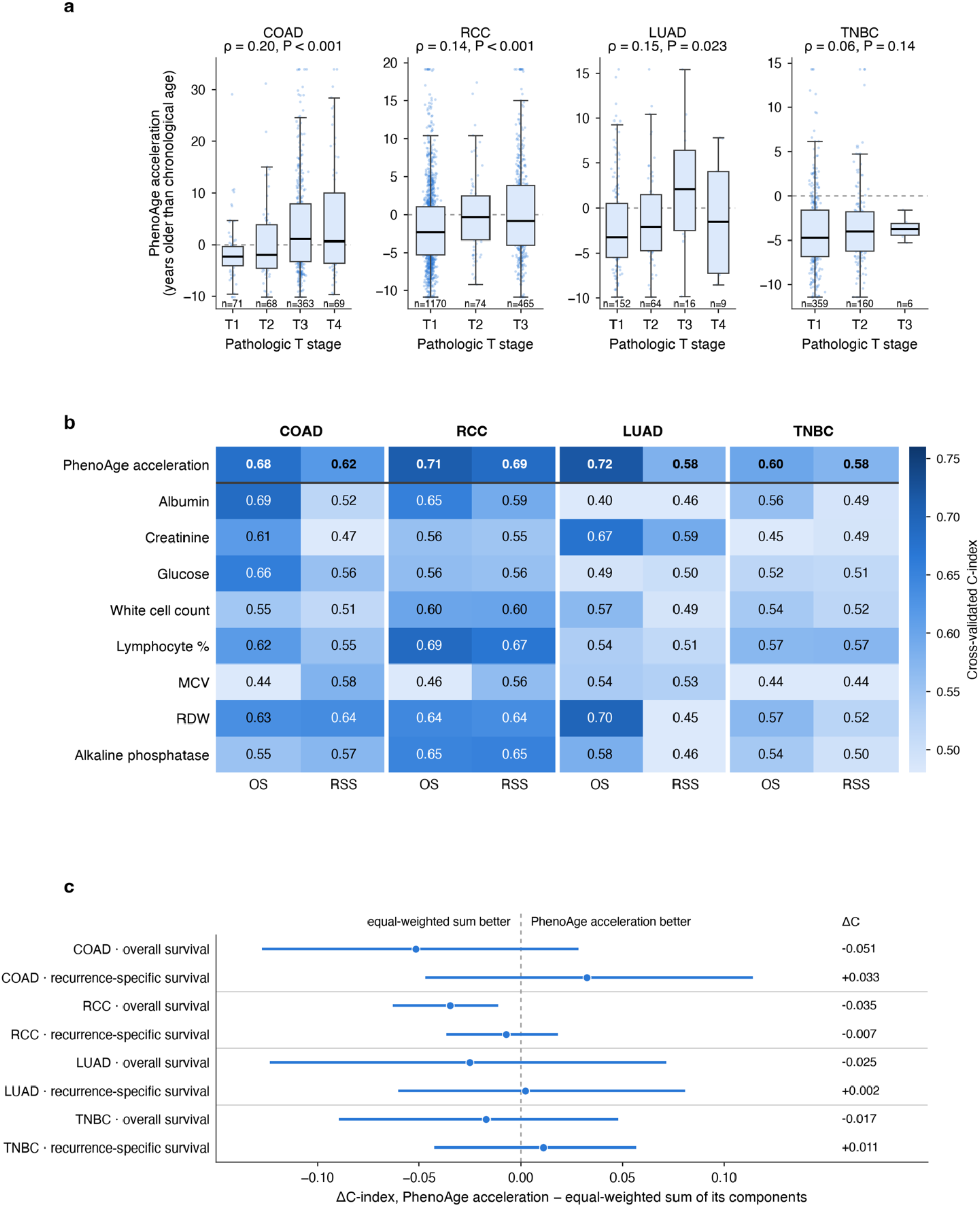
PhenoAge acceleration tracks tumor burden, its signal is carried by cancer-specific components and its weights are not prognostic. (a) PhenoAge acceleration, in years older than chronological age, by pathologic T stage, one panel per cancer. Boxes are the median and interquartile range, whiskers the furthest values within 1.5 times the interquartile range and each dot a patient, with values outside the cohort’s 1st to 99th percentile drawn at that limit; the number of patients at each stage is given beneath each box. ρ is the Spearman correlation between acceleration and T stage. COAD and RCC use the 571 and 1,709 patients with an ordinal T category. In RCC and TNBC, the T3 box includes T4. (b) Cross-validated C-index of each PhenoAge component alone, with PhenoAge acceleration as the reference row, for overall survival (OS) and recurrence-specific survival (RSS) in each cancer. Each cell is one Cox model of that component, scored by 15 repeats of 5-fold cross-validation; values near or below 0.50 mean the component carries no usable signal. (c) Difference in cross-validated C-index between PhenoAge acceleration and an equal-weighted sum of all eight components, in which each PhenoAge coefficient is replaced by its sign; points are the mean paired difference and bars its 95% interval over 500 out-of-bag bootstrap resamples. Negative values favor the equal-weighted sum. C-index, concordance index; COAD, colon adenocarcinoma; LUAD, lung adenocarcinoma; MCV, mean corpuscular volume; OS, overall survival; RCC, renal cell carcinoma; RDW, red cell distribution width; RSS, recurrence-specific survival; TNBC, triple-negative breast cancer.

We then asked whether acceleration improves on each cancer’s clinicopathologic model. Each cancer’s established clinicopathologic predictors reached a C-index for overall survival of 0.79 in COAD, 0.80 in RCC, 0.74 in LUAD and 0.64 in TNBC. Adding acceleration raised it by 0.013 to 0.023, with the bootstrap interval excluding zero only in RCC; for recurrence-specific survival the increments were 0.000 to 0.017 and no interval excluded zero (Supplementary Table S3). A measure of underlying biological aging would not be expected to track tumor stage in these patients undergoing curative resection, and it should add prognostic information beyond established clinicopathologic factors.

### The prognostic signal is carried by one or two cancer-specific routine components

In each cancer type, we scored each component alone by the same cross-validated procedure used for acceleration (Fig. 2b, Supplementary Table S3). In COAD a single component, albumin, matched acceleration (C-index 0.69 versus 0.68); in RCC a single component, lymphocyte percentage, nearly matched it (0.69 versus 0.71). In LUAD two components carried the signal: red cell distribution width (RDW; 0.70) and creatinine (0.67); no other reached 0.59. In TNBC no component reached 0.58; lymphocyte percentage, RDW and albumin were within 0.01 of one another, against 0.60 for acceleration. In each cancer type, adding a second component to the strongest closed the remaining gap. A pair built on each cancer’s leading component had a C-index no more than 0.01 below acceleration’s in every cancer: albumin and glucose (0.72) in COAD, albumin and lymphocyte percentage (0.71) in RCC, creatinine and RDW (0.71) in LUAD, lymphocyte percentage and RDW (0.60) in TNBC. In COAD eight of the 28 possible pairs exceeded acceleration; in LUAD and TNBC two or three pairs were interchangeable at the top, so the named pair is representative rather than unique. Tertiles of the pair separated overall-survival curves as tertiles of acceleration did (Fig. 1, right column). The same held once the clinical predictors were in the model: the pair raised the clinical C-index by 0.028 in COAD, 0.024 in RCC and 0.036 in LUAD, against 0.013, 0.023 and 0.013 for acceleration, and by 0.008 in TNBC against 0.014. Every interval for the pair included zero; in LUAD, RDW alone added 0.042 (Supplementary Table S3). For recurrence-specific survival the pattern held but the component changed: RDW in COAD (0.64 versus 0.62 for acceleration), lymphocyte percentage in RCC (0.67 versus 0.69), creatinine in LUAD (0.59 versus 0.58) and lymphocyte percentage in TNBC (0.57 versus 0.58). Albumin, lymphocyte percentage, RDW and creatinine are the values oncologists already read as nutritional depletion, immune suppression, marrow stress and renal or cachectic compromise.

### PhenoAge’s weights are not prognostic in cancer

PhenoAge weights each component by a coefficient estimated to predict mortality in the U.S. general population (NHANES), and these coefficients were held fixed here, as in standard works on PhenoAge and cancer. To test whether they carry prognostic information in cancer, we replaced every coefficient by its sign alone (+1 or −1) and added the eight standardized components with no age term. This equal-weighted sum had a C-index at least as high as acceleration in every cancer for overall survival (0.73 versus 0.68 in COAD, 0.74 versus 0.71 in RCC, 0.73 versus 0.72 in LUAD, 0.62 versus 0.60 in TNBC) and within 0.03 of it for recurrence-specific survival (Fig. 2c, Supplementary Table S3). The bootstrap interval for the difference excluded zero only for overall survival in RCC, where the equal-weighted sum was higher. In cancer, PhenoAge’s value lies in combining components that already carry prognostic meaning. The weights, which are essential for its function as a biological clock, add nothing to prognosis in these cohorts.

Across four biologically distinct cancers, PhenoAge predicted outcome. Yet PhenoAge acceleration rose with tumor stage and added little to established clinicopathologic factors. Furthermore, its association with survival was carried by one or two routine components that differed by cancer and by endpoint, and this was reproduced when its population-derived weights were replaced by their signs. This pattern is not what a general measure of aging would show. A simpler explanation is that PhenoAge is sensitive to the systemic consequences of malignancy, which are already reflected in the routine blood tests it happens to contain.

This study has several limitations. It is retrospective, single-center and restricted to surgical patients with a complete pre-operative panel. We used a modified eight-component PhenoAge because CRP was rarely measured, although the nine-component formula, with the missing CRP handled in two ways, gave similar results on both endpoints (Supplementary Table S2). None of the models were tested in an independent cohort from another institution. Finally, stage and laboratory abnormalities are imperfect measures of disease burden, and we had no independent measure of biological aging. Our findings therefore cannot establish that PhenoAge contains no aging signal in patients with cancer, only that much of its prognostic information here is reproduced by cancer-associated laboratory abnormalities.

Components such as RDW, albumin, lymphocyte percentage and alkaline phosphatase reflect inflammation, nutritional status, immune function and organ or tumor burden, all known to influence cancer outcomes. PhenoAge integrates these abnormalities, weights them according to a reference population, and expresses them as a “biological age.” An association between accelerated PhenoAge and poor cancer outcomes can invite mechanistic conclusions about aging that PhenoAge itself does not establish. Our findings do not imply that biological aging is irrelevant to cancer; different clocks measure different things^19^, and we tested only one. Rather, our findings suggest that studying aging in oncology will require measures whose signal can be distinguished from the inflammatory, nutritional, hematologic and organ-specific consequences of the cancer and its treatment.

## Methods

### Cohorts and patient selection

We analyzed 3,067 patients who underwent curative-intent resection of a primary solid tumor at Memorial Sloan Kettering Cancer Center: 577 with colon adenocarcinoma (COAD), 1,724 with renal cell carcinoma (RCC), 241 with lung adenocarcinoma (LUAD) and 525 with triple-negative breast cancer (TNBC). Each group was drawn from a larger source cohort assembled for a previously published onco-anesthesia study: COAD (n=1,157)^13^, RCC (n=2,735)^14^, LUAD (n=740)^13^ and TNBC (n=1,143)^15^. The study was approved by the Memorial Sloan Kettering Institutional Review Board (protocol 18-538). A patient was included in this study if all eight PhenoAge components were present in a single laboratory panel drawn before the day of surgery and no more than one year earlier. Patients given systemic therapy before surgery were excluded, so that the measured PhenoAge was not confounded by previous treatment. A complete panel was available for 577 COAD, 1,724 RCC, 293 LUAD and 755 TNBC patients. No COAD or RCC patient had received pre-operative systemic therapy; 52 LUAD patients (51 given induction chemotherapy and one whose treatment status could not be linked) and 230 TNBC patients given neoadjuvant chemotherapy were excluded. Blood was drawn a median of 7, 10, 14 and 7 days before surgery in COAD, RCC, LUAD and TNBC, respectively. Baseline characteristics are in Supplementary Table S1. All RCC patients were clinically non-metastatic at presentation; 18 in whom pathologic M1 disease was identified at surgery were retained.

### Endpoints

Overall survival (OS) was death from any cause. Recurrence-specific survival (RSS) took each cohort’s recorded recurrence as the event (first recurrence in COAD, development of metastasis in RCC and recurrence in LUAD and TNBC) and censored patients who died without one. Time was measured from the date of surgery and censored at last follow-up. Only the Kaplan–Meier curves in Fig. 1 and their log-rank tests were censored at five years.

### PhenoAge

PhenoAge was computed from its published formulation^16^, in which a Gompertz proportional-hazards linear predictor built from chronological age and its blood-based components is transformed into an age in years. Because C-reactive protein (CRP) is rarely measured before surgery, the coefficients were re-estimated on NHANES III with the CRP term omitted, following the BioAge procedure^17^ as in a previous surgical cohort study^18^. All eight retained components kept the sign of their original coefficients (Supplementary Table S2). In the one cohort with measured CRP (RCC, n=211), the eight- and nine-component formulations ranked patients almost identically (Spearman ρ = 0.99); that subset has too few deaths for a separate survival analysis. In all four cohorts the nine-component formula was also applied with the missing CRP held at the NHANES population mean or with the term dropped, and each formulation was evaluated by the same cross-validation on both endpoints. Rankings were near-identical (ρ > 0.96) and C-indexes were within 0.02 in every cohort (Supplementary Table S2). PhenoAge acceleration is PhenoAge minus chronological age and was analyzed as a continuous variable throughout, as was each component value.

### Comparison with components

Within each cancer, we compared a Cox model of PhenoAge acceleration with a Cox model of (1) each of its eight components alone, (2) a two-component model built on the most prognostic component, and (3) an equal-weighted sum of all eight. The two-component model for each cancer was the pair with the highest cross-validated C-index for overall survival among the 28 possible pairs; it was chosen on all patients and carried unchanged to recurrence-specific survival. In LUAD and TNBC, multiple pairs discriminated equally well, so the pair shown is representative.

### Equal-weighted sum

The equal-weighted sum replaces each PhenoAge coefficient by its sign (+1 or −1), adds the eight components standardized within the training fold with no age term, and is scored as a single covariate. PhenoAge’s coefficients were estimated in NHANES and are fixed constants here, so this tests whether the weights themselves carry information. These comparisons included no clinical covariates, since PhenoAge and its components derive from the same pre-operative panel and the question is whether the composite carries information its components do not.

### Cross-validation and bootstrap

Out-of-sample discrimination was estimated by 5-fold cross-validation stratified on the event indicator, with covariates standardized within each training fold and a ridge-penalized Cox model (penalty 0.05) scored on the held-out fold. The C-index reported for each model is the mean over 15 repeats with different random partitions; a single partition moved the C-index by up to 0.07 in LUAD, where there were 22 deaths, so the repeats are needed for the comparisons to be stable. Intervals came from a separate procedure: 500 out-of-bag bootstrap resamples, in each of which every model was fitted on the in-bag patients and scored on those not drawn. The 95% interval is the 2.5th to 97.5th percentile of the paired differences in C-index computed on the same resamples (Supplementary Table S3). Intervals containing zero were interpreted as inconclusive evidence of a difference, not as evidence of equivalence.

### Survival curves

Patients were divided into tertiles of PhenoAge acceleration and, separately, of the two-component composite, defined as the linear predictor of a Cox model of that cancer’s representative pair fitted on the full cohort. Tertiles of PhenoAge acceleration were formed on its value directly; for a single covariate these coincide with tertiles of its Cox linear predictor. The curves are descriptive and censored at five years so that the four cancers share one axis.

### Clinical models

For each cancer, PhenoAge acceleration and, separately, that cancer’s leading component and its representative pair (as in Fig. 1) were added to a ridge-penalized Cox model of the clinicopathologic factors associated with outcome in the source studies. The COAD model contained age, pathologic T and N stage, histologic grade, mismatch-repair status and the van Walraven comorbidity index. The RCC model contained age, T and N stage, histology category, nuclear grade (low, 1–2, versus high, 3–4) and the van Walraven comorbidity index. The LUAD model contained age, pathologic stage group, grade, lepidic versus other histology and American Society of Anesthesiologists (ASA) class. The TNBC model contained age, locoregional treatment, histology, lymphovascular invasion, T and N stage, adjuvant therapy and the van Walraven index. Categorical covariates were expanded to indicators, with unknown as a level. Incremental value was the change in cross-validated C-index, with its out-of-bag bootstrap interval, on both endpoints (Supplementary Table S3).

### Tumor stage and hazard ratios

PhenoAge acceleration was correlated (Spearman) with ordinal pathologic T stage within each cancer (COAD Tis and RCC TX excluded). Per-SD hazard ratios for PhenoAge acceleration on both endpoints were estimated with unpenalized Cox models, unadjusted; for overall survival the model was refitted with T stage as an ordinal covariate.

### Software and reproducibility

PhenoAge coefficients were re-estimated following the BioAge procedure^17^; all other analyses were performed in Python using lifelines, scikit-survival, scikit-learn, pandas, NumPy, SciPy and statsmodels. Code will be available on publication at github.com/aksimhal/phenoage.

## Data Availability

Data produced in the present study are available upon reasonable request to the authors in accordance with MSK policies.

## Acknowledgements

JSM acknowledges Sahrena London for helpful conversations.

## References

1. Liu, Z. et al. A new aging measure captures morbidity and mortality risk across diverse subpopulations from NHANES IV: A cohort study. PLoS Med. 15, e1002718 (2018).

2. Liu, Z. et al. Correction: A new aging measure captures morbidity and mortality risk across diverse subpopulations from NHANES IV: A cohort study. PLoS Med. 16, e1002760 (2019).

3. Tian, R. et al. Biological aging and generational shifts in early-onset cancer risk. Nat. Med. 32, 2983–2990 (2026).

4. Yang, J., Zhang, Y., Zeng, P. & Jian, X. PhenoAgeAccel mediates the association between metabolic syndrome and cancer risk in males: a cross-sectional NHANES study. Aging Male 29, 2612796 (2026).

5. Isogai, N. et al. Association between lumbar spinal stenosis and accelerated biological aging estimated by PhenoAge. J. Clin. Med. 14, 7852 (2025).

6. Xi, J. et al. PhenoAgeAccel is associated with all-cause and cardiovascular mortality in patients with diabetes and prediabetes: A cohort study. Diabetes Res. Clin. Pract. 226, 112375 (2025).

7. Bian, L. et al. Associations of combined phenotypic aging and genetic risk with incident cancer: A prospective cohort study. Elife 13, (2024).

8. Wang, X. et al. Joint association of biological aging and lifestyle with risks of cancer incidence and mortality: A cohort study in the UK Biobank. Prev. Med. 182, 107928 (2024).

9. McMillan, D. C. Systemic inflammation, nutritional status and survival in patients with cancer. Curr. Opin. Clin. Nutr. Metab. Care 12, 223–226 (2009).

10. Diakos, C. I., Charles, K. A., McMillan, D. C. & Clarke, S. J. Cancer-related inflammation and treatment effectiveness. Lancet Oncol. 15, e493–503 (2014).

11. Yuval, J. B. et al. Intraoperative opioids are associated with decreased recurrence rates in colon adenocarcinoma: a retrospective observational cohort study. Br. J. Anaesth. 129, 172–181 (2022).

12. Silagy, A. W. et al. Impact of intraoperative opioid and adjunct analgesic use on renal cell carcinoma recurrence: role for onco-anaesthesia. Br. J. Anaesth. 125, e402–e404 (2020).

13. Connolly, J. G. et al. Intraoperative opioid exposure, tumour genomic alterations, and survival differences in people with lung adenocarcinoma. Br. J. Anaesth. 127, 75–84 (2021).

14. Montagna, G. et al. Intraoperative opioids are associated with improved recurrence-free survival in triple-negative breast cancer. Br. J. Anaesth. 126, 367–376 (2021).

15. Liu, W. et al. Oxidative stress factors mediate the association between Life’s Essential 8 and accelerated phenotypic aging: NHANES 2005-2018. J. Gerontol. A Biol. Sci. Med. Sci. 79, glad240 (2024).

16. Luo, L., Fu, J., Wang, X. & Sui, X. The association between phenotypic age acceleration and the risk of all-cause and cancer mortality among cancer survivors: NHANES 1999-2018. Sci. Rep. 16, 1349 (2025).

17. Kwon, D. & Belsky, D. W. A toolkit for quantification of biological age from blood chemistry and organ function test data: BioAge. GeroScience 43, 2795–2808 (2021).

18. Scarpa, J. R., Reid, M. C., Elemento, O. & Krumsiek, J. Biological aging increases risk of postoperative morbidity and mortality: an international, multi-cohort study. NPJ Aging 10.1038/s41514-026-00441-2 (2026) doi:10.1038/s41514-026-00441-2.

19. Belsky, D. W. et al. Eleven telomere, epigenetic clock, and biomarker-composite quantifications of biological aging: Do they measure the same thing? Am. J. Epidemiol. 187, 1220–1230 (2018).

