## Supplemental Table for "Blood-based PhenoAge may reflect disease burden rather than biological aging in cancer"

### Supplementary Tables S1–S3

Blood-based PhenoAge may reflect disease burden rather than biological aging in cancer

### Table S1. Baseline characteristics of the four surgical cohorts

| Characteristic | Unit / summary | Colon (COAD) | Kidney (RCC) | Lung (LUAD) | Breast (TNBC) |
|---|---|---|---|---|---|
| **Cohort** | | | | | |
| N | n | 577 | 1,724 | 241 | 525 |
| **Demographics** | | | | | |
| Age at surgery | yr, median [IQR] | 60.6 [50.8, 71.1] | 60.5 [52.6, 68.2] | 67.8 [62.1, 73.3] | 55.1 [45.8, 64.5] |
| Female sex | n (%) | 291 (50) | 568 (33) | 157 (65) | 525 (100) |
| **Race** | **n (%)** | | | | |
| White | | 479 (83) | 1454 (84) | 192 (80) | 375 (71) |
| Black | | 30 (5) | 100 (6) | 10 (4) | 81 (15) |
| Asian / Pacific Islander | | 41 (7) | 78 (5) | 26 (11) | 40 (8) |
| Other | | 8 (1) | 35 (2) | 0 (0) | 11 (2) |
| Unknown / declined | | 0 (0) | 57 (3) | 13 (5) | 0 (0) |
| Missing / not recorded | | 19 (3) | 0 (0) | 0 (0) | 18 (3) |
| Hispanic or Latino ethnicity | n (%) | 34 (6) | 98 (6) | 3 (1) | N/A |
| Body mass index | kg/m², median [IQR] | 27.7 [24.3, 32.4] | 29.3 [26.1, 33.4] | 26.2 [23.3, 29.2] | 26 [23, 31] |
| **Comorbidity** | | | | | |
| van Walraven comorbidity index | score, median [IQR] | 12 [8, 12] | 4 [4, 12] | N/A | 4 [4, 9] |
| **ASA physical status** | **n (%)** | | | | |
| I–II | | 209 (36) | 545 (32) | 29 (12) | 310 (59) |
| III–IV | | 368 (64) | 1179 (68) | 212 (88) | 215 (41) |
| **Tumor** | | | | | |
| **Pathologic T stage** | **n (%)** | | | | |
| Tis | | 6 (1) | N/A | 0 (0) | 0 (0) |
| T1 | | 71 (12) | 1170 (68) | 152 (63) | 359 (68) |
| T2 | | 68 (12) | 74 (4) | 64 (27) | 160 (30) |
| T3–4 | | 432 (75) | 465 (27) | 25 (10) | 6 (1) |
| TX | | 0 (0) | 15 (1) | 0 (0) | 0 (0) |
| **Pathologic N stage** | **n (%)** | | | | |
| N0 | | 382 (66) | 500 (29) | 192 (80) | 389 (74) |
| NX | | 0 (0) | 1189 (69) | 0 (0) | 0 (0) |
| N+ | | 195 (34) | 35 (2) | 49 (20) | 136 (26) |
| **Grade** | **n (%)** | | | | |

| | | | | | |
|---|---|---|---|---|---|
| Well / moderately differentiated | | 448 (78) | N/A | 117 (49) | 33 (6) |
| Poorly differentiated | | 115 (20) | N/A | 62 (26) | 492 (94) |
| Low (1–2) | | N/A | 651 (38) | N/A | N/A |
| High (3–4) | | N/A | 754 (44) | N/A | N/A |
| Missing / not recorded | | 14 (2) | 319 (19) | 62 (26) | 0 (0) |
| **Pre-operative blood draw** | | | | | |
| Days from draw to surgery | d, median [IQR] | 7 [5, 13] | 10 [6, 17] | 14 [8, 24] | 7 [6, 12] |
| Draw > 30 d before surgery | n (%) | 10 (2) | 137 (8) | 42 (17) | 10 (2) |
| **PhenoAge components*** | | | | | |
| Albumin | g/dL | 4.2 [4.0, 4.4] | 4.3 [4.2, 4.5] | 4.2 [4.0, 4.4] | 4.3 [4.2, 4.5] |
| Creatinine | mg/dL | 0.90 [0.70, 1.00] | 1.00 [0.80, 1.10] | 0.80 [0.80, 1.00] | 0.80 [0.70, 0.90] |
| Glucose | mg/dL | 100 [87, 120] | 97 [86, 115] | 100 [91, 117] | 94 [83, 110] |
| Lymphocyte percentage | % | 23.5 [19.0, 28.3] | 24.6 [19.3, 30.1] | 26.5 [20.9, 32.2] | 29.5 [23.7, 34.3] |
| Mean corpuscular volume | fL | 86.0 [80.0, 90.0] | 88.0 [85.0, 91.0] | 90.0 [87.0, 93.0] | 90.0 [86.0, 93.0] |
| Red cell distribution width | % | 13.9 [12.9, 16.4] | 13.2 [12.7, 13.9] | 13.2 [12.7, 14.0] | 13.0 [12.6, 13.6] |
| Alkaline phosphatase | U/L | 74 [62, 91] | 75 [62, 91] | 74 [60, 86] | 73 [59, 91] |
| White blood cell count | $\times 10^3/\mu L$ | 6.9 [5.7, 8.0] | 6.7 [5.7, 8.0] | 6.9 [5.8, 8.2] | 6.3 [5.3, 7.6] |
| C-reactive protein measured | n (%) | 1 (0) | 211 (12) | 0 (0) | 0 (0) |
| C-reactive protein (where measured) | mg/dL, median [IQR] | N/A | 0.23 [0.11, 0.53] | N/A | N/A |
| **PhenoAge** | | | | | |
| PhenoAge | yr, median [IQR] | 61.8 [50.2, 77.1] | 59.3 [49.9, 68.6] | 66.1 [59.1, 72.9] | 51.0 [42.0, 60.9] |
| PhenoAge acceleration | yr, median [IQR] | 0.0 [-3.7, 6.9] | -1.9 [-4.9, 1.8] | -2.7 [-5.1, 1.5] | -4.4 [-6.6, -1.7] |
| PhenoAge acceleration > 0 | n (%) | 287 (50) | 616 (36) | 80 (33) | 79 (15) |

*Shown in clinical units; values are converted to the model units of Table S2 before PhenoAge is computed.

1. Continuous variables are median [IQR], with the IQR given as [Q1, Q3]; categorical variables are n (%), with the cohort N as the denominator throughout, so a variable's levels and its Missing / not recorded row sum to 100%; that row is omitted where the variable is complete in every cohort. N/A marks a cell with no number to report, either because that cohort did not collect the variable or because the category does not exist for that cancer (RCC has no Tis; differentiation and nuclear grade are separate scales). 0 (0) means the category applies but no patient had it. ASA, American Society of Anesthesiologists; IQR, interquartile range; N/A, not applicable.

**Table S2. The eight-component and nine-component PhenoAge formulations: coefficients, agreement and the handling of missing CRP**

| (a) Term | Units | Eight-component | Nine-component |
|---|---|---|---|
| **Gompertz linear predictor (xb)** | | | |
| Intercept | — | -17.770519284 | -19.9067 |
| Albumin | g/L | -0.028894491 | -0.0336 |
| Creatinine | µmol/L | 0.0068392 | 0.0095 |
| Glucose | mmol/L | 0.104152143 | 0.1953 |
| ln(C-reactive protein) | ln(mg/dL) | not in model | 0.0954 |
| Lymphocyte percentage | % | -0.011517186 | -0.012 |
| Mean corpuscular volume | fL | 0.024864997 | 0.0268 |
| Red cell distribution width | % | 0.241783074 | 0.3306 |
| Alkaline phosphatase | U/L | 0.002104687 | 0.00188 |
| White blood cell count | $\times 10^3/\mu L$ | 0.061915138 | 0.0554 |
| Chronological age | years | 0.077707238 | 0.0804 |
| **Mortality-to-age transform** | | | |
| γ (Gompertz shape) | — | 0.007459589 | 0.0076927 |
| A (age intercept) | years | 83.602451 | 141.50225 |
| c (scale inside the log) | — | 1 | 0.00553 |
| S (age slope) | — | 0.085489137 | 0.090165 |

(a) The eight-component formulation is the BioAge “phenoage_calc” model refitted in NHANES III without CRP and is the primary model throughout; the nine-component formulation is the original published model, shown for comparison. Components enter in the model units shown; clinical values are converted before evaluation.

| (b) Quantity | Value |
|---|---|
| Patients with a measured CRP (RCC) | 211 |
| Pearson r, nine-component versus eight-component PhenoAge | 0.986 |
| Spearman ρ, nine-component versus eight-component PhenoAge | 0.987 |
| Spearman ρ, nine-component versus eight-component PhenoAge acceleration | 0.973 |
| Median difference, nine minus eight (SD), years | -1.7 (2.8) |
| Deaths in this subset | 9 |
| Composite recurrence-free survival, 12 events: apparent C, nine / eight | 0.698 / 0.685 |
| cross-validated C, nine / eight | 0.697 / 0.678 |
| bootstrap difference, nine minus eight (95% CI) | +0.011 (-0.018 to +0.055) |

(b) The one cohort with a measured CRP is RCC (n=211 of 1,724). In these patients the published nine-component PhenoAge, computed with the measured CRP, and the eight-component refit used throughout rank patients almost identically, so the eight-component PhenoAge stands in for the nine-component one; the nine-component PhenoAge is a median 1.7 years lower. The subset has too few deaths for a survival comparison, and the composite recurrence-free survival comparison shown rests on 12 events and is not informative on its own.

C-index, concordance index; CI, confidence interval; COAD, colon adenocarcinoma; CRP, C-reactive protein; LUAD, lung adenocarcinoma; NHANES, National Health and Nutrition Examination Survey; OS, overall survival; RCC, renal cell carcinoma; RDW, red cell distribution width; RSS, recurrence-specific survival; SD, standard deviation; TNBC, triple-negative breast cancer.

| (c) Cancer | Endpoint | n (events) | ρ, nine-component (CRP at mean) versus primary | Shift, years | C, primary eight-component | C, nine-component, CRP at mean | C, nine-component, CRP term dropped | C, representative pair |
|---|---|---|---|---|---|---|---|---|
| COAD | OS | 577 (52) | 0.989 | +0.8 | 0.679 | 0.683 | 0.683 | 0.725 (Albumin + glucose) |
| COAD | RSS | 576 (62) | 0.989 | +0.8 | 0.618 | 0.620 | 0.620 | 0.536 (Albumin + glucose) |
| RCC | OS | 1,724 (132) | 0.982 | -0.4 | 0.710 | 0.691 | 0.691 | 0.708 (Albumin + lymphocyte %) |
| RCC | RSS | 1,724 (117) | 0.982 | -0.4 | 0.685 | 0.672 | 0.672 | 0.662 (Albumin + lymphocyte %) |
| LUAD | OS | 241 (22) | 0.978 | -0.6 | 0.718 | 0.708 | 0.708 | 0.714 (Creatinine + RDW) |
| LUAD | RSS | 241 (53) | 0.978 | -0.6 | 0.580 | 0.575 | 0.575 | 0.569 (Creatinine + RDW) |
| TNBC | OS | 525 (69) | 0.969 | -1.2 | 0.597 | 0.584 | 0.584 | 0.595 (Lymphocyte % + RDW) |
| TNBC | RSS | 525 (86) | 0.969 | -1.2 | 0.576 | 0.565 | 0.565 | 0.572 (Lymphocyte % + RDW) |

(c) The primary formulation is the eight-component refit with no CRP term. Keeping the published nine-component formula instead and filling the missing CRP with a constant, either the NHANES population mean (0.42 mg/dL) or the value that removes the term (1 mg/dL), gives a PhenoAge that ranks patients almost identically to the primary one (Spearman ρ 0.969 to 0.989) and has a C-index within 0.019 of it in every cohort and endpoint. The two constants differ by an additive shift only, so their C-indexes are identical. The comparison with each cancer's representative pair is unchanged under any formulation. The RCC patients with a measured CRP are in part (b).

Cross-validated C-index as in Table S3 (15 repeats of 5-fold cross-validation, ridge penalty 0.05). ρ and shift compare the nine-component PhenoAge with CRP at the NHANES mean against the primary formulation on the same patients.
C-index, concordance index; CI, confidence interval; COAD, colon adenocarcinoma; CRP, C-reactive protein; LUAD, lung adenocarcinoma; NHANES, National Health and Nutrition Examination Survey; OS, overall survival; RCC, renal cell carcinoma; RDW, red cell distribution width; RSS, recurrence-specific survival; SD, standard deviation; TNBC, triple-negative breast cancer.

**Table S3. Discrimination of PhenoAge acceleration, of each of its components alone, of a representative pair and of an equal-weighted sum, and their value beyond clinical predictors**

**(a) Cross-validated C-index of each model for every cancer and endpoint**

| Model | COAD OS | COAD RSS | RCC OS | RCC RSS | LUAD OS | LUAD RSS | TNBC OS | TNBC RSS |
|---|---|---|---|---|---|---|---|---|
| n (events) | 577 (52) | 576 (62) | 1,724 (132) | 1,724 (117) | 241 (22) | 241 (53) | 525 (69) | 525 (86) |
| PhenoAge acceleration | 0.679 | 0.618 | 0.710 | 0.685 | 0.718 | 0.580 | 0.597 | 0.576 |
| Albumin | 0.686 | 0.520 | 0.654 | 0.590 | 0.402 | 0.455 | 0.558 | 0.489 |
| Creatinine | 0.614 | 0.468 | 0.557 | 0.549 | 0.674 | 0.590 | 0.453 | 0.490 |
| Glucose | 0.658 | 0.556 | 0.563 | 0.557 | 0.491 | 0.503 | 0.518 | 0.513 |
| WBC | 0.547 | 0.515 | 0.599 | 0.601 | 0.568 | 0.491 | 0.543 | 0.519 |
| Lymphocyte % | 0.616 | 0.554 | 0.689 | 0.670 | 0.535 | 0.513 | 0.571 | 0.572 |
| MCV | 0.439 | 0.578 | 0.461 | 0.563 | 0.538 | 0.530 | 0.439 | 0.442 |
| RDW | 0.633 | 0.637 | 0.643 | 0.636 | 0.697 | 0.453 | 0.569 | 0.522 |
| Alkaline phosphatase | 0.552 | 0.571 | 0.649 | 0.652 | 0.583 | 0.461 | 0.543 | 0.496 |
| Representative pair* | 0.725 | 0.536 | 0.708 | 0.662 | 0.714 | 0.569 | 0.595 | 0.572 |
| Equal-weighted sum of all eight | 0.730 | 0.588 | 0.745 | 0.693 | 0.734 | 0.576 | 0.616 | 0.566 |

**(b) Paired differences from PhenoAge acceleration, with 95% out-of-bag bootstrap intervals**

| Cancer | Endpoint | Leading component | PhenoAge acceleration − leading component (95% CI) | PhenoAge acceleration − pair (95% CI) | PhenoAge acceleration − equal-weighted sum (95% CI) |
|---|---|---|---|---|---|
| COAD | OS | Albumin | -0.008 (-0.135 to +0.122) | -0.042 (-0.143 to +0.067) | -0.051 (-0.127 to +0.028) |
| COAD | RSS | Albumin | +0.110 (-0.029 to +0.329) | +0.099 (-0.024 to +0.322) | +0.033 (-0.047 to +0.114) |
| RCC | OS | Lymphocyte % | +0.022 (-0.055 to +0.089) | +0.002 (-0.066 to +0.071) | -0.035 (-0.063 to -0.011) |
| RCC | RSS | Lymphocyte % | +0.017 (-0.064 to +0.084) | +0.024 (-0.047 to +0.101) | -0.007 (-0.037 to +0.018) |
| LUAD | OS | RDW | +0.023 (-0.097 to +0.148) | +0.005 (-0.132 to +0.166) | -0.025 (-0.123 to +0.072) |
| LUAD | RSS | RDW | +0.094 (-0.094 to +0.262) | +0.009 (-0.154 to +0.195) | +0.002 (-0.060 to +0.081) |
| TNBC | OS | RDW | +0.036 (-0.078 to +0.221) | +0.010 (-0.092 to +0.123) | -0.017 (-0.089 to +0.048) |
| TNBC | RSS | RDW | +0.065 (-0.038 to +0.222) | +0.010 (-0.094 to +0.120) | +0.011 (-0.043 to +0.057) |

**(c) Cross-validated C-index of each cancer's clinical model alone and with each addition; in parentheses, the change over the clinical model with its 95% interval**

| Cancer | Endpoint | n (events) | Clinical model | + PhenoAge acceleration | + leading component | + pair |
|---|---|---|---|---|---|---|
| COAD | OS | 577 (52) | 0.785 | 0.798 (+0.013; -0.033 to +0.054) | 0.801 (+0.015; -0.033 to +0.055) | 0.813 (+0.028; -0.023 to +0.075) |
| COAD | RSS | 576 (62) | 0.736 | 0.737 (+0.000; -0.037 to +0.023) | 0.734 (-0.003; -0.035 to +0.012) | 0.731 (-0.005; -0.040 to +0.013) |
| RCC | OS | 1,724 (132) | 0.802 | 0.824 (+0.023; +0.001 to +0.045) | 0.819 (+0.017; -0.009 to +0.035) | 0.826 (+0.024; -0.012 to +0.051) |
| RCC | RSS | 1,724 (117) | 0.853 | 0.870 (+0.017; -0.000 to +0.031) | 0.865 (+0.012; -0.010 to +0.027) | 0.867 (+0.014; -0.014 to +0.033) |
| LUAD | OS | 241 (22) | 0.738 | 0.751 (+0.013; -0.046 to +0.061) | 0.780 (+0.042; -0.050 to +0.140) | 0.774 (+0.036; -0.058 to +0.144) |
| LUAD | RSS | 241 (53) | 0.752 | 0.754 (+0.002; -0.038 to +0.032) | 0.745 (-0.007; -0.042 to +0.011) | 0.752 (-0.000; -0.053 to +0.036) |
| TNBC | OS | 525 (69) | 0.636 | 0.650 (+0.014; -0.011 to +0.049) | 0.641 (+0.005; -0.029 to +0.030) | 0.645 (+0.008; -0.035 to +0.045) |
| TNBC | RSS | 525 (86) | 0.661 | 0.668 (+0.007; -0.015 to +0.027) | 0.660 (-0.001; -0.024 to +0.014) | 0.663 (+0.002; -0.034 to +0.027) |

(a) Every model is a ridge-penalized Cox model (penalty 0.05) scored by 15 repeats of 5-fold cross-validation stratified on the event indicator, with covariates standardized within each training fold; the C-index is the mean over the 75 held-out folds. Each component row is one Cox model of that component alone. The equal-weighted sum adds the eight standardized components, each multiplied by the sign of its PhenoAge coefficient, with no age term, and is scored as one covariate. *Representative pairs, selected on overall survival and carried unchanged to recurrence-specific survival, are albumin + glucose (COAD), albumin +

lymphocyte % (RCC), creatinine + RDW (LUAD) and lymphocyte % + RDW (TNBC). Values near or below 0.50 mean the component carries no usable signal for that endpoint.
(b) The leading component is the one with the highest C-index for overall survival in that cancer, carried unchanged to recurrence-specific survival. Differences are PhenoAge acceleration minus the comparison model, computed over 500 out-of-bag bootstrap resamples in which every model was fitted on the in-bag patients and scored on those not drawn, and summarized by the 2.5th and 97.5th percentiles; a negative difference favors the comparison model. An interval containing zero is inconclusive evidence of a difference, not evidence of equivalence.
(c) Clinical models contain the clinicopathologic factors of each source study (Methods), assembled for overall survival and used unchanged for recurrence-specific survival. The value in parentheses is the change in cross-validated C-index over the clinical model, with its 95% interval from 500 out-of-bag bootstrap resamples.
C-index, concordance index; CI, confidence interval; COAD, colon adenocarcinoma; LUAD, lung adenocarcinoma; MCV, mean corpuscular volume; OS, overall survival; RCC, renal cell carcinoma; RDW, red cell distribution width; RSS, recurrence-specific survival; TNBC, triple-negative breast cancer; WBC, white blood cell count.
